# Whole Genome Sequencing Informed Patient Personalized Measurable Residual Disease Assays for Acute Myeloid Leukemia

**DOI:** 10.64898/2026.01.22.26343677

**Authors:** Niveditha Ravindra, Justin Lack, Clifton L. Dalgard, Eddy vanCollenburg, Adam Corner, Lan Beppu, Harry Erba, Megan Othus, Jerald P. Radich, Laura W. Dillon, Christopher S. Hourigan

## Abstract

Post-treatment measurable residual disease (MRD) in acute myeloid leukemia (AML) patients is associated with adverse clinical outcomes. Validated molecular methods for AML MRD are preferable to flow cytometry assays but are not available for all patients. The limit of detection (LOD) of next-generation sequencing (NGS) assays for single nucleotide variants is restricted by technical error rates. Structural alterations are common genetic features of AML, but MRD approaches for detecting this class of variants have primarily relied on RNA. However, RNA has suboptimal stability, not all structural alterations are expressed as transcripts, and the impact of anti-leukemic therapy on transcription may make leukemic disease burden quantification inaccurate. In this study, we demonstrate a whole genome sequencing (WGS)-based approach to identify genomic DNA breakpoints of chromosomal rearrangements that allowed design of highly sensitive patient-personalized digital droplet PCR (ddPCR) MRD assays.

Acute myeloid leukemia (AML) is an aggressive malignancy of the hematopoietic precursor cells that predominantly affects older individuals. Oncogenic transformation occurring through the acquisition of structural chromosomal aberrations is noted in 35% of AML cases, and can result in the formation of fusion proteins that confer proliferation and survival advantages (1). When compared to classical cytogenetics for the identification of structural variants at diagnosis, newer techniques such as optical genome mapping can identify clinically pertinent aberrations that may be cryptic or smaller than the resolution of conventional karyotyping and FISH (2). Similarly, short-read whole genome sequencing (WGS) has been shown to increase diagnostic yield and better refine risk stratification when compared to traditional cytogenetic testing in myeloid malignancies (3). Additionally, WGS can be utilized to identify genomic breakpoints of chromosomal rearrangements at a basepair (bp) resolution.

## Main Text

The presence of measurable residual disease (MRD) in complete remission (CR) is prognostic of outcomes in patients with AML (4) and validated molecular methods for MRD monitoring are preferable to flow cytometry assays but are not available for all patients (5). The limit of detection (LOD) of next-generation sequencing (NGS) assays for single nucleotide variants is restricted by technical error rates and not all patients have a molecular variant detectable by standard NGS assays. Structural alterations are common genetic features of AML, but MRD approaches for detecting this class of variants have primarily relied on RNA. However, RNA has suboptimal stability, not all structural alterations are expressed as transcripts, and the impact of anti-leukemic therapy on transcription may make leukemic disease burden quantification inaccurate. These shortcomings could largely be overcome by using DNA-based assays which provide accurate quantification of MRD and prevent contamination, although breakpoints are widely distributed in intronic regions (6). In this study, we describe a sensitive DNA-based patient-personalized approach to MRD detection in AML by droplet digital PCR (ddPCR) using breakpoint information from WGS.

Bone marrow (BM) or peripheral blood samples from 62 adult patients with AML enrolled on the randomized Phase 3 SWOG-S0106 trial (NCT00085709) were previously assessed by our group for MRD in first CR by multiparametric flow cytometry (MFC) and duplex sequencing (DS) (7). While MRD detection by DS was highly predictive of adverse outcomes and strongly outperformed MFC, it did not predict relapse in all cases and 8% of patients did not have an appropriate variant available for monitoring. Therefore, we sought to investigate if WGS-informed MRD tracking of structural alterations could overcome this limitation by selecting 5 patients from this cohort who experienced relapse and had known cytogenetic abnormalities detected at diagnosis (**Table 1**), consisting of 2 patients without a DS-trackable variant and 3 patients with DS-trackable variants (*FLT3, NRAS*, and *KIT*). Among selected patients with trackable variants, there were 2 false negatives and 1 true positive for MRD by DS. The Institutional Review Board of the Fred Hutchinson Cancer Center gave ethical approval for this work, and patients were treated according to the Declaration of Helsinki. Full details on experimental methods are outlined in the supplement.

**Table 1:**
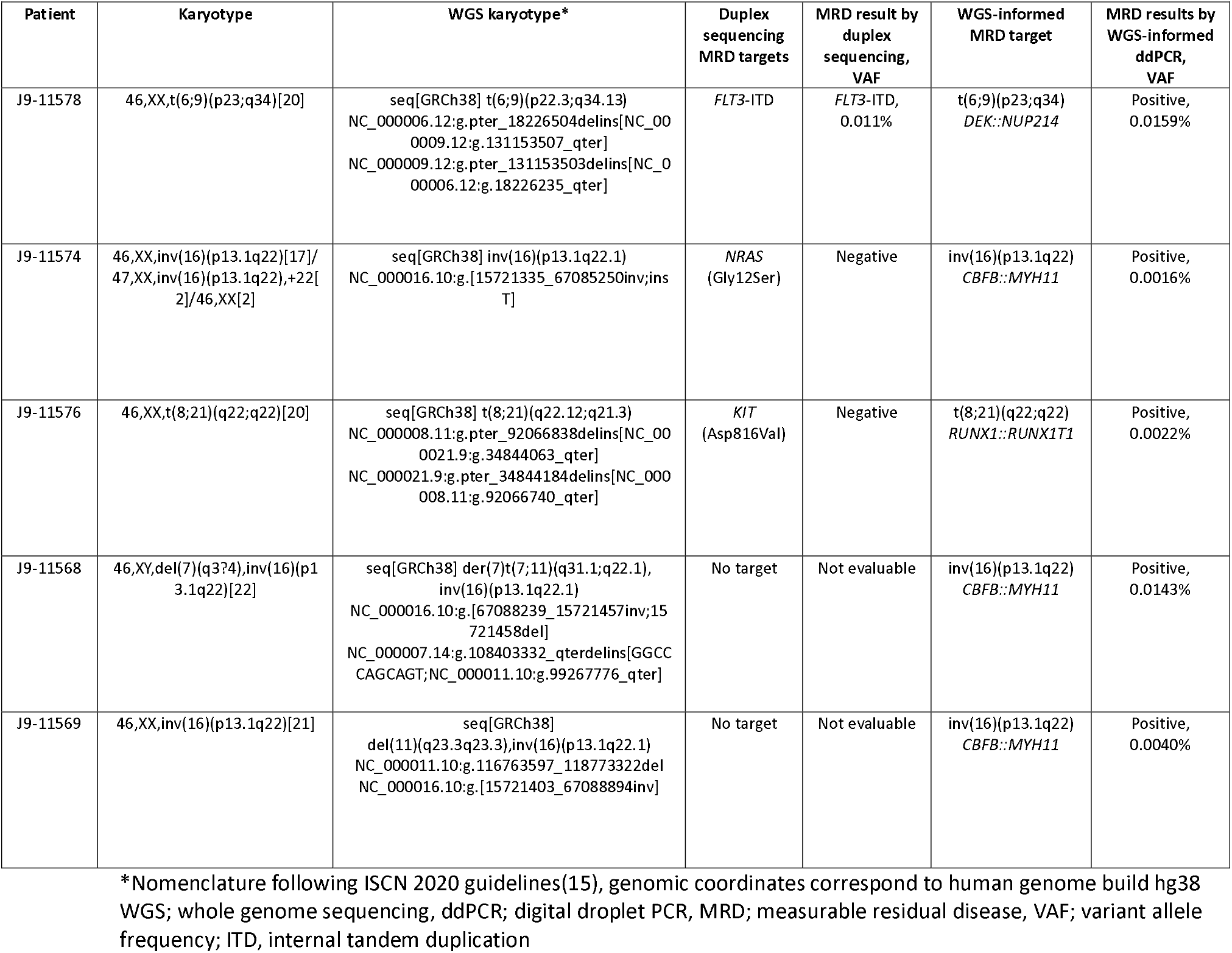
Comparison of conventional and WGS-based karyotyping at baseline and MRD by duplex sequencing and WGS-informed ddPCR.

| Patient | Karyotype | WGS karyotype* | Duplex sequencing MRD targets | MRD result by duplex sequencing, VAF | WGS-informed MRD target | MRD results by WGS-informed ddPCR, VAF |
| --- | --- | --- | --- | --- | --- | --- |
| J9-11578 | 46,XX,t(6;9)(p23;q34)[20] | seq[GRCh38] t(6;9)(p22.3;q34.13)<br>NC_000006.12:g.pter_18226504delins[NC_000009.12:g.131153507_qter]<br>NC_000009.12:g.pter_131153503delins[NC_000006.12:g.18226235_qter] | *FLT3*-ITD | *FLT3*-ITD, 0.011% | t(6;9)(p23;q34)<br>*DEK::NUP214* | Positive, 0.0159% |
| J9-11574 | 46,XX,inv(16)(p13.1q22)[17]/47,XX,inv(16)(p13.1q22),+22[2]/46,XX[2] | seq[GRCh38] inv(16)(p13.1q22.1)<br>NC_000016.10:g.[15721335_67085250]inv;ins T] | *NRAS*<br>(Gly12Ser) | Negative | inv(16)(p13.1q22)<br>*CBFB::MYH11* | Positive, 0.0016% |
| J9-11576 | 46,XX,t(8;21)(q22;q22)[20] | seq[GRCh38] t(8;21)(q22.12;q21.3)<br>NC_000008.11:g.pter_92066838delins[NC_000021.9:g.34844063_qter]<br>NC_000021.9:g.pter_34844184delins[NC_000008.11:g.92066740_qter] | *KIT*<br>(Asp816Val) | Negative | t(8;21)(q22;q22)<br>*RUNX1::RUNX1T1* | Positive, 0.0022% |
| J9-11568 | 46,XY,del(7)(q37.4),inv(16)(p13.1q22)[22] | seq[GRCh38] der(7)t(7;11)(q31.1;q22.1),<br>inv(16)(p13.1q22.1)<br>NC_000016.10:g.[67088239_15721457]inv;15721458del]<br>NC_000007.14:g.108403332_qterdelins[GGCC CAGCAGT;NC_000011.10:g.99267776_qter] | No target | Not evaluable | inv(16)(p13.1q22)<br>*CBFB::MYH11* | Positive, 0.0143% |
| J9-11569 | 46,XX,inv(16)(p13.1q22)[21] | seq[GRCh38]<br>del(11)(q23.3q23.3),inv(16)(p13.1q22.1)<br>NC_000011.10:g.116763597_118773322del<br>NC_000016.10:g.[15721403_67088894]inv] | No target | Not evaluable | inv(16)(p13.1q22)<br>*CBFB::MYH11* | Positive, 0.0040% |

\*Nomenclature following ISCN 2020 guidelines(15), genomic coordinates correspond to human genome build hg38 WGS; whole genome sequencing, ddPCR; digital droplet PCR, MRD; measurable residual disease, VAF; variant allele frequency; ITD, internal tandem duplication

Initially, we performed a proof of concept experiment by designing a ddPCR assay for the genomic *CBFB::MYH11* breakpoints previously characterized in the ME-1 cell line (8). For all ddPCR analysis performed in this study, protocols suggested by the manufacturer were followed (Bio-Rad Laboratories Inc., Hercules, CA). Briefly, 300-900ng (300ng/reaction) of genomic DNA was combined with ddPCR supermix for probes (no dUTP) (Catalog number: 1863025, Bio-Rad Laboratories Inc., Hercules, CA, USA), HindIII restriction enzyme (Catalog number: R0140S, New England BioLabs Inc., Ipswich, CA, USA), and mutant and reference gene primers and probes at a final concentration of 900 nM and 250 nM, respectively. This was followed by droplet generation, PCR, droplet reading, and data analysis using the QuantaSoft software. Serial dilution of ME-1 cell line DNA into healthy donor DNA (range 5%-0.001%) established the LOD of the cell line-specific *CBFB::MYH11* assay as 0.001% variant allele frequency (VAF), with high sensitivity, no background noise, and high correlation to the expected VAF (r ^2^ =0.99, P<0.0001) (**Supplementary Figure 1**).

For the 5 AML patients, WGS libraries were constructed from baseline BM DNA using the TruSeq DNA PCR-free library preparation kit (Illumina, San Diego, California, USA) and subjected to 150-bp paired-end WGS to an average depth of 65X on the NovaSeq 6000 platform (Illumina) according to the manufacturer’s protocols. The analytic pipeline for structural variants (SVs) and copy-number changes (CNVs) was modified from the Chromoseq pipeline described by Duncavage *et al* (3). Specifically, this workflow flags and retains recurrent and risk-defining SVs and CNVs while more stringent filtering criteria are applied to novel variants. The workflow of the study has been outlined in **Figure 1A**.

**Figure 1.**
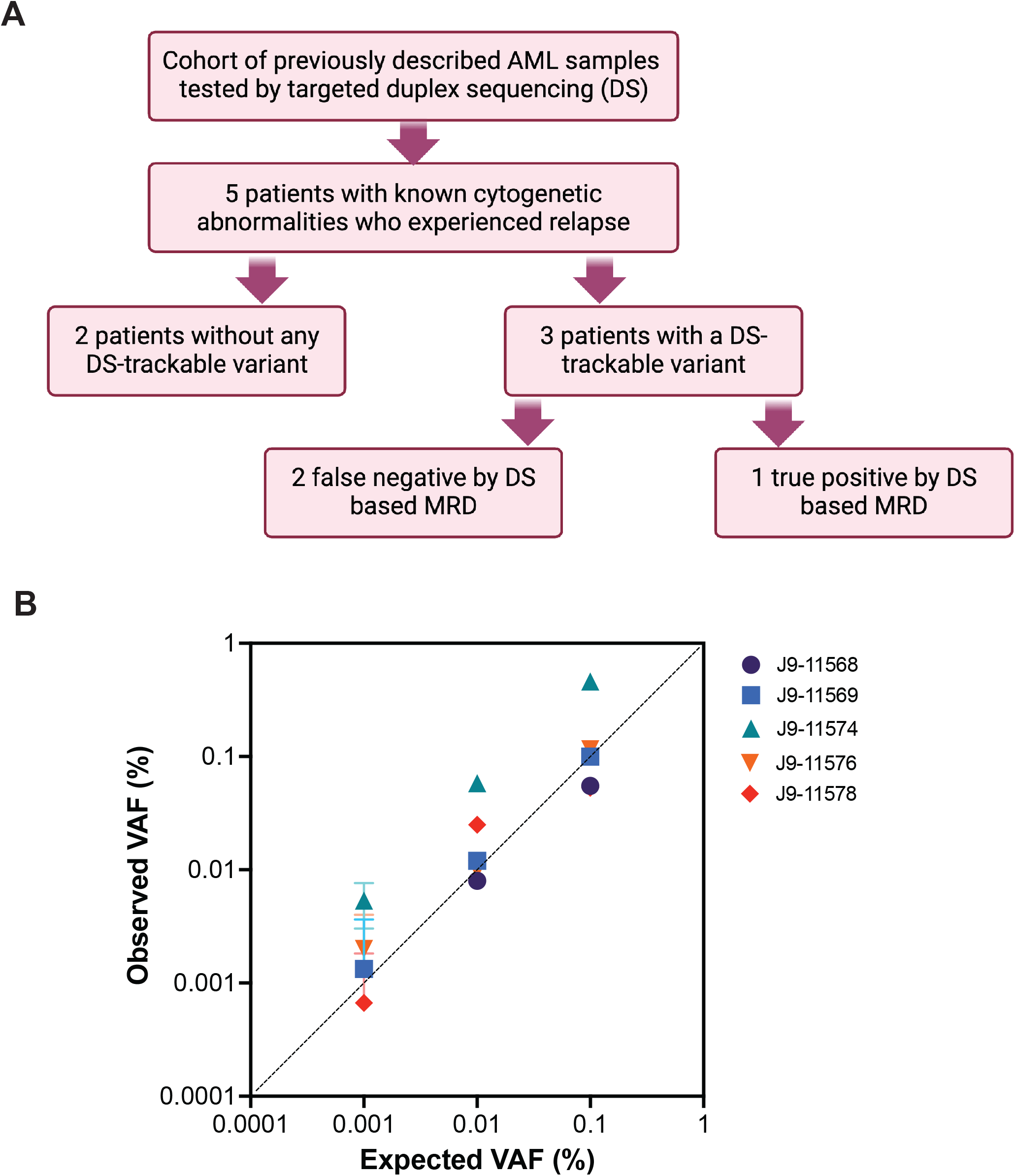
Assessment of whole-genome sequencing-informed patient personalized digital PCR assays. (A) Outline of the study design. (B) Expected versus observed variant allele fraction (VAF) of patient-personalized whole genome sequencing-informed digital PCR assays performed on serial dilutions of positive control DNA. A limit of detection for each assay was achieved ranging anywhere from 0.01%-0.001%.

WGS successfully identified the genomic DNA breakpoints of all anticipated chromosomal rearrangements in the 5 diagnostic AML samples (**Table 1**). In one patient (J9-11568), WGS was able to resolve an unclear cytogenetic finding on the q arm of chromosome (chr) 7 as a derivative chr resulting from a translocation between chr 7 and 11, der(7)t(7;11)(q31.1;q22.1), resulting in a 50Mb net loss of material on 7q31.1-q36.3 and 35Mb gain of material on 11q22.1-q25. In a second patient (J9-11569), WGS identified a 2Mb deletion on 11q23.3. In both cases, the patient had core-binding factor AML harboring inv(16), and the identification of secondary cytogenetic abnormalities within this disease class has been linked to distinct prognostic impacts depending on the abnormalities identified (9).

One structural event was selected per patient and the breakpoints were confirmed by Sanger sequencing, including *CBFB*::*MYH11* (n=3), *RUNX1*::*RUNX1T1* (n=1), and *DEK*::*NUP214* (n=1) (**Table 1, Supplementary Table 1**). Custom FAM-tagged ddPCR assays targeting the translocation event were designed for each patient and multiplexed with a HEX-tagged *EIF2C1* reference gene assay (Bio-Rad Laboratories Inc., Hercules, CA, assay ID: dHsaCP1000002) for calculation of wild-type genome copies. Individual assays were validated using 900ng of input DNA by serial dilution experiments (range 1%-0.0001%), establishing LODs ranging from 0.01%-0.001% VAF (**Figure 1B**).

Patient-personalized assays were subsequently tested on post-treatment remission BM samples and identified MRD positivity in all 5 patients who relapsed, with VAFs ranging from 0.0016%-0.016% (**Table 1**). This contrasted with DS, which predicted relapse in a single patient, and MFC which was negative for MRD in all patients. Patient J9-11578, who was MRD positive by both molecular methods had similar residual disease levels by each technique, with a VAF of 0.016% for the *DEK::NUP214* translocation by WGS-informed ddPCR and 0.011% for *FLT3*-ITD by DS. The remaining two patients with DS-trackable variants had residual SV VAFs of 0.0016% and 0.0022% for the *CBFB::MYH11* and *RUNX1::RUNX1T1* fusions, respectively.

This study demonstrates a sensitive, patient-personalized strategy for AML MRD detection for chromosomal structural variants that uses breakpoint information from WGS and DNA as the analyte. This approach was able to successfully predict relapse in all remission samples in the small cohort of patients tested and highlights the sensitivity and specificity of targeting unique chromosomal breakpoints as compared to other more technically error-prone variants such as SNVs. When comparing RNA- and DNA-based methods for AML MRD detection for common fusions, Lukes *et al* showed that genomic DNA-based assays were more sensitive than RNA-based assays, and low transcript levels at diagnosis significantly affected MRD detection using RNA at later time points (10). In another study performed on a cohort of pediatric AML patients, genomic MRD positivity at day 28 was the only significant predictor for overall and event-free survival by multivariate analysis (11). Both studies used targeted next generation (NGS) with custom probes capturing the fusions of interest. However, WGS can provide additional information regarding the diagnostic karyotype, such as chromosomal gains and losses in addition to genomic breakpoints. Moreover, copy number changes such as deletions of important tumor suppressor genes and passenger variants can also be potential MRD targets, which will require future investigation. MRD detection by NGS is gaining wider acceptance, especially for *FLT3*-ITD and *NPM1* which are highly prognostic for relapse (12), and it would be reasonable to spike-in primers/probes targeting SVs into multigene NGS panels.

While WGS-based MRD monitoring offers several benefits, the major factors limiting its use in clinical laboratories would be the cost and expertise required to implement bioinformatics workflows, interpretation of results, and the regulatory burden of validating patient-specific assays. Furthermore, given the sensitivity of WGS (13, 14), tumor cell enrichment in AMLs with low blast percentages and/or deep WGS may be required. While the workflow described in this study requires more extensive validation in larger patient cohorts to establish clinical utility and predictive MRD levels, it provides the opportunity to expand applicability of MRD testing to all AML patients.

## Data Availability

All data produced in the present work are contained in the manuscript.

## Acknowledgments and Funding

This work was funded, in part, by the Intramural Research Program of the National Heart, Lung, and Blood Institute (NHLBI). Sequencing was performed at the American Genome Center, Uniformed Services University of the Health Sciences. Digital droplet PCR experiments were performed with the support of the Center for Cancer Research Genomics core at the National Cancer Institute. Support for the FNIH Biomarkers Consortium MRD in AML (Measurable Residual Disease in Acute Myeloid Leukemia) project was provided by AbbVie; Amgen; AstraZeneca; Genentech, a member of the Roche Group; Gilead Sciences, Inc.; GSK; Jazz Pharmaceuticals, Inc.; LGC Clinical Diagnostics, Inc.; Novartis; Syndax Pharmaceuticals, Inc.; Sysmex Corportation. In-kind donations of standards and methods materials and equipment to support the project were provided to the FNIH by AccuGenomics, Inc.; Bio-Rad Laboratories, Inc.; Invivoscribe, Inc.; LGC Clinical Diagnostics, Inc.; Mission Bio; 10x Genomics, Inc.; Takeda Pharmaceuticals U.S.A., Inc.; Thermo Fisher Scientific Inc.; TwinStrand Biosciences, Inc.; Twist Bioscience Corporation.

The views expressed in this article do not reflect the official policy or position of the National Institute of Health, the Department of the Navy, the Department of Defense, Health Resources and Services Administration (HRSA) or any other agency of the U.S. Government.

