## Supplementary Appendix for "Whole Genome Sequencing Informed Patient Personalized Measurable Residual Disease Assays for Acute Myeloid Leukemia"

### Supplementary data

### Methods

#### 1. Patients and samples

Bone marrow (BM) or peripheral blood (PB) samples from 62 patients with acute myeloid leukemia (AML) enrolled in the randomized Phase 3 SWOG-S0106 trial (NCT00085709) were previously assessed for measurable residual disease (MRD) in the first complete remission using multiparametric flow cytometry and a duplex sequencing (DS) panel targeting 29 genes. From this cohort, we selected 5 patients who experienced relapse and had known cytogenetic abnormalities at diagnosis: 2 patients without a DS-trackable variant and 3 patients with a DS-trackable variant (2 false negative and 1 true positive). All patients included provided written and informed consent. The Institutional Review Board of the Fred Hutchinson Cancer Center gave ethical approval for this work, and patients were treated according to the Declaration of Helsinki.

#### 2. Whole Genome Sequencing (WGS)

Genomes of baseline acute myeloid leukemia (AML) were sequenced according to the manufacturer's protocols. Briefly, 1ug of DNA was used for preparing libraries using the Illumina TruSeq DNA PCR-free library preparation kit (San Diego, California 92122, USA). DNA was mechanically fragmented to produce insert sizes of 350 base pairs (bp) on the Covaris M220 instrument. Fragmented DNA was cleaned up using sample purification (SP) beads which is a component of the kit, followed by end repair, size selection, adenylation of the 3' ends, and ligation of adapters (Illumina DNA Single Index adapters, Set A (San Diego, California 92122, USA). Ligated DNA was subjected to two rounds of purification using SP beads. Quality control of the sequencing-ready libraries was performed on the Agilent Tapestation to confirm the library size distribution and quantified using the KAPA Library Quant Kit for Illumina (KAPA Biosystems, Cape Town, South Africa), to determine loading conditions for sequencing. Libraries were sequenced to a depth of 65X (paired-end, 150 bp) on the Illumina NovaSeq 6000 platform.

### 3. Bioinformatics analysis

For genomic characterization of AML, we implemented the Chromoseq pipeline to efficiently identify clinically relevant structural variants in a tumor-only workflow [1]. For initial processing, raw reads were trimmed with fastp v0.23.4, mapped to the GRCh38 human reference genome using bwa-mem2, and duplicate reads were marked using samblaster v0.1.26 [2, 3]. For the Chromoseq workflow, structural variants (SVs) >=100 kbp in length were called using Manta v1.6.0 and copy number variants (CNVs) > 5Mbp were called using Canvas v1.40.0 [4, 5]. Duplications and deletions were then annotated with breakpoint log2 fold changes (coverage within the breakpoints vs outside of breakpoints) relative to flanking regions using Duphold v0.2.3 [6]. Known SVs and CNVs previously identified as recurrent or risk-defining (list curated by Duncavage et al.) are flagged and retained, while novel events underwent more stringent filtering [1].

For novel SVs to be retained in the final call set, they had to pass the following filters: 1) at least 2 paired and 2 split reads supporting the break-ends, 2) does not overlap blacklist set of recurrent SVs identified by Abel et al. [7] and provided as part of the Chromoseq package (https://github.com/genome/docker-basespace_chromoseq), 3) coverage depth ratio <0.8 or >1.3 relative to background for deletions or duplications, respectively, 4) the breakpoint-spanning contigs must map back to the reported breakpoints. The resulting CNVs and SVs for a given sample are then merged using jasmine v1.1.5 and annotated using VEP 106 (McLaren et al. 2016) and AnnotSV v3.3.1 [8], [9], [10].

### 4. Sanger Sequencing

Breakpoints identified by WGS were confirmed by Sanger sequencing. Primers were designed flanking the intronic breakpoints of the chromosomal translocation. The sequence of interest was amplified using the Q5 high-fidelity PCR kit (New England Biolabs). The amplified product was purified using SPRISelect beads (Beckman Coulter). Sequencing reactions were set up with the BigDye Terminator v3.1 Cycle Sequencing kit (ThermoFisher Scientific) and capillary electrophoresis was performed on the ABI Genetic Analyzer 3500 (Applied Biosystems).

### 5. Digital droplet polymerase chain reaction (ddPCR)

Personalized ddPCR assays for measurable residual disease (MRD) based on breakpoint information from WGS were designed for each patient in this study. Primers were designed flanking the breakpoints of the translocation to amplify sequences ranging from 77-99 bp. Taqman probe sequences were designed within the amplicon and were tagged with the FAM fluorophore. The HEX-tagged *EIF2C1* gene assay (dHsaCP1000002, Bio-Rad Laboratories Inc., Hercules, CA, USA) was used to calculate reference genome copies in all assays.

DNA was combined with ddPCR supermix for probes (no dUTP) (Catalog number: 1863025, Bio-Rad Laboratories Inc., Hercules, CA, USA), HindIII restriction enzyme (Catalog number: R0140S, New England BioLabs Inc., Ipswich, CA, USA), along with mutant and reference gene primers and probes at a final concentration of 900 nM and 250 nM respectively. 20uL of the mix was carried forward for droplet generation using the Automated Droplet Generator (Bio-Rad Laboratories Inc., Hercules, CA, USA) to generate ~20,000 droplets per reaction. PCR was performed using the following conditions: Initial denaturation at 95°C for 10 minutes was followed by 40 cycles each of 94°C for 30 seconds and 55°C for 1 minute with an enzyme deactivation step of 98°C for 1 minute. Droplet fluorescence was analyzed on the QX200 Droplet Reader (cat # 18640003, Bio-Rad Laboratories, Inc., Hercules, CA). Data analysis was carried out using the QuantaSoft Analysis Pro ver. 1.0.596 software (Bio-Rad Laboratories, Inc., Hercules, CA). Wells with fewer than 10,000 droplets were excluded from the analysis. Thresholds separating positive and negative droplets were set using negative controls.

Due to the limited availability of starting material, validation of individual assays was performed using PCR products generated for confirmation by Sanger sequencing. Amplicons were diluted in healthy donor DNA to simulate variant allele frequencies (VAFs) ranging from

0.1% to 0.0001%. Triplicates of 300ng/well of each dilution were run along with an adequate number of negative controls to calculate the limit of detection (LOD) for each assay.

Following validation, the remission samples were tested using a total input of 900ng of DNA distributed in 3 wells. Negative and no-template controls were included in each run. Samples were considered positive if fluorescence was detected above the LOD for the corresponding assay.

**Supplementary Figure 1. Serial dilution of ME-1 cell line.**
DNA from ME-1 cell line was diluted into DNA from a healthy donor at an expected variant allele fraction (VAF) of 5%-0.001% and the level of *CBFB::MYH11* was assessed at the DNA level using an ddPCR assay targeting the DNA breakpoints. The anticipated versus observed VAF is plotted with a line of equivalence plotted as a dashed line. The Pearson correlation and p-value are reported on the inset of the graph.

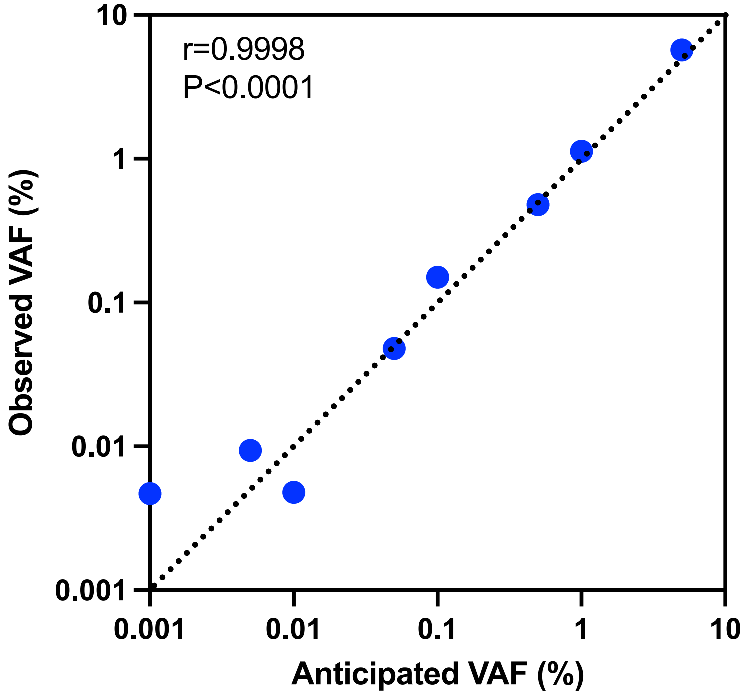

**Supplementary Table 1. Sanger sequencing validation of genomic DNA breakpoints identified by whole genome sequencing.**

| Patient | WGS breakpoint | Sanger sequencing confirmed DNA breakpoint sequence |
|---|---|---|
| J9-11578 | 6:18226235/9:131153503 | TAATTCATTCCAGACTCTCAGCCTGAAAACACTGAGAATGTTTGCATGCTAGTTTTCCACATCATATACAATATTATTAAAAT<br>ACTCATTTGGAATAGAATTCCATATGGGTTAACCAGAGTACTGTTGGGATGGTTGTGGCTATTTGCACGTAGCAGATTTCCT<br>GCTTTTATTCAAAGACAATATTACTGGATTTTAAAATCTGCTTTTAACATTATTTTTCCTTTTCACTATACATAGGTCTATGAAA<br>ATTACAGGCATCCGCTCTGCATCCGAGCTACATGTTGTTCCTATGTGCCTTTCCACCTCTTTCTCTATTGACAAACTAAGAGGA<br>AAGCATACAGCTTTAGGGAGACACTTATTCAGGAGGGAATAATTTTCTTTTTTCATGGTGCTAAGTAGATAATACTAGTTAGC<br>ATGTATTGAGTGTTTACTATGGACAATGCACAGATGATGAAACCAAGATAACTGAAACTGCCCAGGTTTTACCACTAATAAAC<br>CTTGAAACCTTGAACCTGGG |
| J9-11574 | 16:15721335/16:67085250 | GAGTGGTGATAGGAATGAAAAAGGCCACCCGACCTCCCTCTGCTGGCCTCCCCGGCAGCACGCACCTGTCTCTGCAGTTGC<br>CTCCTCTTCTCCTCATTCTGCTCGTCCCGGGCTTGGAGATCCCTTTCGAACTGGCCCTTGAGCGCCTGCATGTTGACTTCCAG<br>CCGCAGTTTGGCGTCCTCCGTGGCTTGCAGCTCGTCCTCCAGCTCTTCCAGCTGCGTCTTCATCTCCTCCATCTGGGTCTCCA<br>GGGCCCGCTTGGACTTCTCCAGCTCATGGACCTGCCGGCAGAGCGGGCAGCCCCATTCTATGAGGCTCAACTTCATGAAGA<br>GATTGAGAAACCCACCGTGAGCGGCACCTCAGGAGATCAGGGAGGTGGCTTTGGCCTCCCACAGGATGCATGGCCGGGAC<br>TCAAGATGACCCCTGAGAGTTCAGACCCCAGCCTTATCCTCGGACCCCCCAACTCAGACCCATCCTCGACTGCCATTCTCAGC<br>CCCTCCCAGCCCCTGCACCAGTCCAAAAACCTCCTTCCATTTCCGATGATAGTTCGCTATGAAAAAGGCC**T**TTGCGGTGAGCC<br>GACATTGTGCCACTGCACTCCAGCCTGGGCGACAGAATGAGACTCCATCACACACACACACAGAGACACACACAGAGACAC<br>ACACACACACACACAATTTATTTCATACACTAACCACCAAATAGGCTTGA |
| J9-11576 | 8:92066838/21:34844063 | TTGGGCTAAGGGAGATGGACCTCTCTTTTGGATGATTTCCTAACTTCTCATCACTTTGACAGTCAATTAAATAGATACAACAC<br>CTAATAAAAATGTACATTAATTTGTTTCTGCCACTGAAAGCCATCTGAAACTGTATTAAATAAAAGCCATAATAGATTGTCCC<br>CTGACAAAGATAAATCCAATTATACTTTGTCTGCAAAAAAACACAAATGGTTTATGTTTTAGTTAGGCTACAATGTAAGATAC<br>AGCTAACCACAAGAAATAAGTGGGATAGAGGGGGAAAGAAAAAGTTAAGGTCTTATCTCAAGGAAACCCCCAGTTACAGC<br>GTAACTGAAAAGTAACCAGAGAGTGGTAACCACCAGTGACATGGATTTGTTGATCAGAGTAGCTCCTGCCCAGGGACAGAC<br>TCTAAAGAGAAGGCGACTTGCAGAA |
| J9-11568 | 16:67088240/16:15721461 | TCTATTCCAGGCAAAACGGTTCTTAATCTCTCCTGGCAGATTTTATTTTTCAGCTTAAGCATCTTTACGGGTGGTGTGGAAATTA<br>ATTTCCCAGCGCTAAGCTGTATTTGTGCAGGCCTCCTGGTTAGGAAACTGGTTTAAATACTCACTGTTTCTTCCATTAACCAAAT<br>ATTTGAGGGAGAGCAAGTCAATTAATTTTTTGGAGACTCTCAGTTTTATCAACTGTAATAATGTTAGATTAGATGATGTTTGAA<br>TCTCTAAAGATAATCTGAATCACAGAGGGGAAACAACATTTTTCTCCAATATTCTCTTTTTAGGAAAAAGAAGGTAAACTGCAA<br>GAAAATAAGGTGACATTAGGAATTTTTTATGGATGGAAAAAAAAGACCATTTCGGCTTTGAGCATTTTGTTGGTCCGCTCGAGT<br>TCCTCTTTGGCTTCCAAGGCCTCTTCAAGGGCCCGAGCCAGGGACAGGGCCTTGGTTTCCTTCTCCCTGGCTTCTGCCTCAGCTC<br>TGTCCCTCTCATCCGCGTATTTGGAAGAGATGTTTTTCTCCTCGGCTAACAACTACAACACAAGACCCAGAGGTGACTTCTAGGC<br>ATATCCGGGGTCAGCGTCACTGAATTGTAAATACCGGGGGAAGCCCTGTGTCCTGCTGAATGTATTGAGGTGCAGGTGTAAGC<br>AGTGTAGGTTAGCTATGGGAGTAATTTATATAATCATCTGGCGTTCTGACTTCTACATCAACCTTATGCAGAGCCA |
| J9-11569 | 16:15721403/16:67088894 | TTCATGAAGACGATTGAGAAACCCACCGTGAGCGGCACCTCAGGAGATCAGGGAGGTGGCTTTGGCCTCCCACAGGATGCAT<br>GGCCGGGACTCAAGATGACCCCTGAGAGTTCAGACCCCAGCCTTATCCTCGGACCCCCCAACTCAGACCCATCCTCGACTGCCA<br>TTCTCAGCCCCTCCCAGCCCCTGCACCAGTCCAAAAACCTCCTTCCATTTCCGATGATAGTTCGCTATGAAAAAGGCCAGGAGCT<br>AGCCTCGCATGGACTGGTGAATAGCACAGAGGGTGGGCAGGCGAAACATGGACGAGAAAAACCCCCTTAGTAATCTTAACCTT<br>ACACATAGATTAAATACTCACGAGCACCACTGTTACAAGTTTAGCATGGCAGGGCTGGAGGCCTCAGAAGTTATCCAAACAGTG<br>GAGGACATCCCTAAACCTCTGTGCTGCTAGGATCTCAAGCTTCCACAGGCCGTTTACTGACTTGCTGCCACCCATGAGTTACCAC<br>TGTTCTCTGGCCCTCCTCTCCAGTCAGGCATCAGGACCACCCACTAGAACCATTATTTTTCTTTTCTGCCTCAGCCTTTTCTATTCC<br>AGGCAAAACGGTTCTTAATC |
